# A 2-year longitudinal investigation of insula subregional volumes in early psychosis

**DOI:** 10.1101/2024.11.25.24317916

**Authors:** Andrew R. Kittleson, Maureen McHugo, Jinyuan Liu, Simon N. Vandekar, Kristan Armstrong, Baxter Rogers, Neil D. Woodward, Stephan Heckers, Julia M. Sheffield

## Abstract

**Background:** The insula is a heterogeneous cortical region with three cytoarchitectural subregions— agranular, dysgranular, and granular—that have distinct functional roles. Previous cross-sectional studies have shown smaller volume of all insula subregions in individuals with psychotic disorders. However, longitudinal trajectories of insula subregions in early psychosis, and the relationship between subregional volumes and relevant clinical phenomena, such as perceptual aberrations, have not been previously examined.

**Methods:** 66 early psychosis (EP) and 65 healthy comparison (HC) participants completed 2-4 study visits over 2 years. T1-weighted structural brain images were processed using longitudinal voxel-based morphometry in CAT12 and segmented into anatomic subregions. At baseline, participants completed the Perceptual Aberrations Scale (PAS) to capture bodily distortions. The EP group was further examined based on diagnostic trajectory over two years (stable schizophrenia, stable schizophreniform, and conversion from schizophreniform to schizophrenia).

**Results:** EP participants had smaller insula volumes in all subregions compared to HC participants, and these volumes were stable over two years. Compared to HC, insula volumes were significantly smaller in EP participants with a stable diagnosis of schizophrenia, but other diagnostic trajectory groups did not significantly differ from HC or the stable schizophrenia group. While perceptual aberrations were significantly elevated in EP participants, PAS scores were not significantly related to insula volume.

**Conclusions:** We find that all insula subregions are smaller in early psychosis and do not significantly decline over two years. These data suggest that all insula subregions are structurally impacted in schizophrenia-spectrum disorders and may be the result of abnormal neurodevelopment.

## Introduction

The human insula is a heterogeneous cortical brain region that is implicated in a wide variety of behaviors. Sometimes referred to as a fifth cerebral lobe, this region is among the first cortical areas to mature and continues to experience rapid growth into the early postnatal period^1,2^. In schizophrenia, insula abnormalities are widely reported, including structural alterations^3^, structural connectivity differences^4^, resting-state functional connectivity changes^5,6^, and abnormal task-based functional activation signatures^7,8^. One of the most common findings in this literature is the presence of smaller insula volumes in schizophrenia, which are observed both in first episode and chronic stages of illness^9^. Insula volumes are also smaller, albeit to a lesser extent, in those at clinical high-risk and with enriched genetic risk for psychosis^10–12^, suggesting that structural changes precede clinical diagnosis.

Yet, the insula itself is not a homogenous region, and can be parsed into more precise subregions based on its cytoarchitecture, anatomy, and associated realms of function^13^. Structural parcellations of the insula often focus on three subregions, distinguished by the relative presence or absence or granular layer IV cortical neurons: agranular (anterior), dysgranular (central), and granular (posterior)^14,15^. These differing neural substrates have implications for associated function^16^. Von Economo neurons (VENs), spatially localized to the agranular insula in cortical layer V, are specialized bipolar neurons that allow for rapid long-range information transfer between brain regions^17,18^. They are thought to facilitate the agranular insula’s roles in complex socio-emotional processing like prosocial behaviors and social empathy^19–22^. Conversely, layer IV granular neurons found densely in the posterior granular insula are smaller and more superficial than VENs. These neurons receive neocortical, subcortical, and even visceral inputs^23–25^, and in synthesizing this sensory information create perceptions of exteroceptive and interoceptive stimuli, implicating the posterior insula in self- embodiment^26,27^. Distinct patterns of subregional volume change over time in early psychosis could reflect selective neuronal degradation in the insula, providing one potential mechanism through which neurobiology affects behavior.

Prior work examining insula volumes in psychotic disorders have largely neglected these subregional distinctions, yet growing evidence points to their relevance. Findings from a large neurodevelopmental sample found smaller insula volumes for the agranular and dysgranular, but not the granular, subregions in psychosis-spectrum youth compared to typically developing youth and youth with other psychopathologies^28^. In addition, individuals with schizophrenia-spectrum disorders, including both those in early and chronic stages, have lower insula volumes compared with nonclinical participants in all subregions. Critically, this volume deficit was not observed in psychotic bipolar disorder, suggesting an additional consideration of diagnostic specificity^28^. Interestingly, the only prior study on longitudinal insula volume in early psychosis reported increases in anterior insula volumes and stable posterior insula volumes in psychosis participants scanned several years after their first-episode of psychosis, using broad anterior/posterior subdivisions and a highly heterogeneous definition of first-episode psychosis^29^. Together, this evidence suggests that insula volume changes in early psychosis may be specific to schizophrenia-spectrum disorders and has some subregional specificity. Yet, how these cytoarchitectural insula subregions change within the early course of illness, in a well-characterized cohort of individuals with schizophrenia-spectrum disorders, remains unknown.

Finally, even within a longitudinal cohort that is robustly characterized and closely followed, schizophrenia-spectrum diagnoses exist along a spectrum of clinical and functional impairment. While some first-episode patients may never meet diagnostic criteria for schizophrenia (e.g., receive and maintain a diagnosis of schizophreniform disorder), others may progress to meet this diagnosis due to persistent or worsening symptoms (e.g., receive a schizophreniform diagnosis during the first episode, but subsequently meet criteria for schizophrenia)^30,31^. With this context, increasing evidence continues to underscore the importance of the insula in the phenomenology of psychotic illness^28,32,33^. With its roles in cognition, emotion, interoception, and salience processing^34,35^, the insula is crucial for a variety of functions that can go awry in psychosis, progressively and throughout the course of illness^36–38^. Indeed, it is possible that individuals with more persistent clinical and functional impairment may demonstrate smaller subregional volumes that decline over the early course of illness. Conversely, if insula alterations are similar across different diagnostic trajectories, this might suggest that insula volume deficits represent a developmental insult that precedes the earliest manifestations of a schizophrenia-spectrum disorder and do not clearly contribute to illness course.

The current study aimed to elucidate insula subregional volume changes in early psychosis. A sample of individuals identified within two years of their first episode for a primary psychotic disorder was followed for two years with structural neuroimaging occurring every 6-8 months. Within this sample, we tested: 1) whether insula subregions demonstrated volumetric differences in early psychosis, 2) how subregional volumes changed over the course of two years in early psychosis participants, and 3) whether diagnostic trajectory subgroups had different patterns of insula volume changes over time. Finally, we explored how insula volumes related to perceptual bodily aberrations, which are linked with insula function, especially in the granular subregion in schizophrenia^39^.

## Methods and Materials

### Study participants

One hundred forty-six (146) total participants (72 healthy comparison [HC] participants and 74 participants in the early stage of a psychotic disorder [EP]) were recruited for a prospective longitudinal study over the early course of psychosis. To target structural pathology unique to the early course of psychotic disorders, EP participants were recruited if it had been less than two years since their first episode of psychosis, with the majority of EP participants recruited within 6-8 months of illness presentation (average duration of psychosis at baseline visit, as measured using Symptom Onset in Schizophrenia Inventory [SOS]^40^, was 6.2 months). EP participants were recruited from outpatient and inpatient settings at Vanderbilt Psychiatric Hospital in Nashville, TN, while HC participants were recruited from the surrounding community. Inclusion and exclusion criteria for both groups can be found in the Supplement. See Table 1 for participant demographics and cognitive characteristics across the entire sample.

**Table 1.**
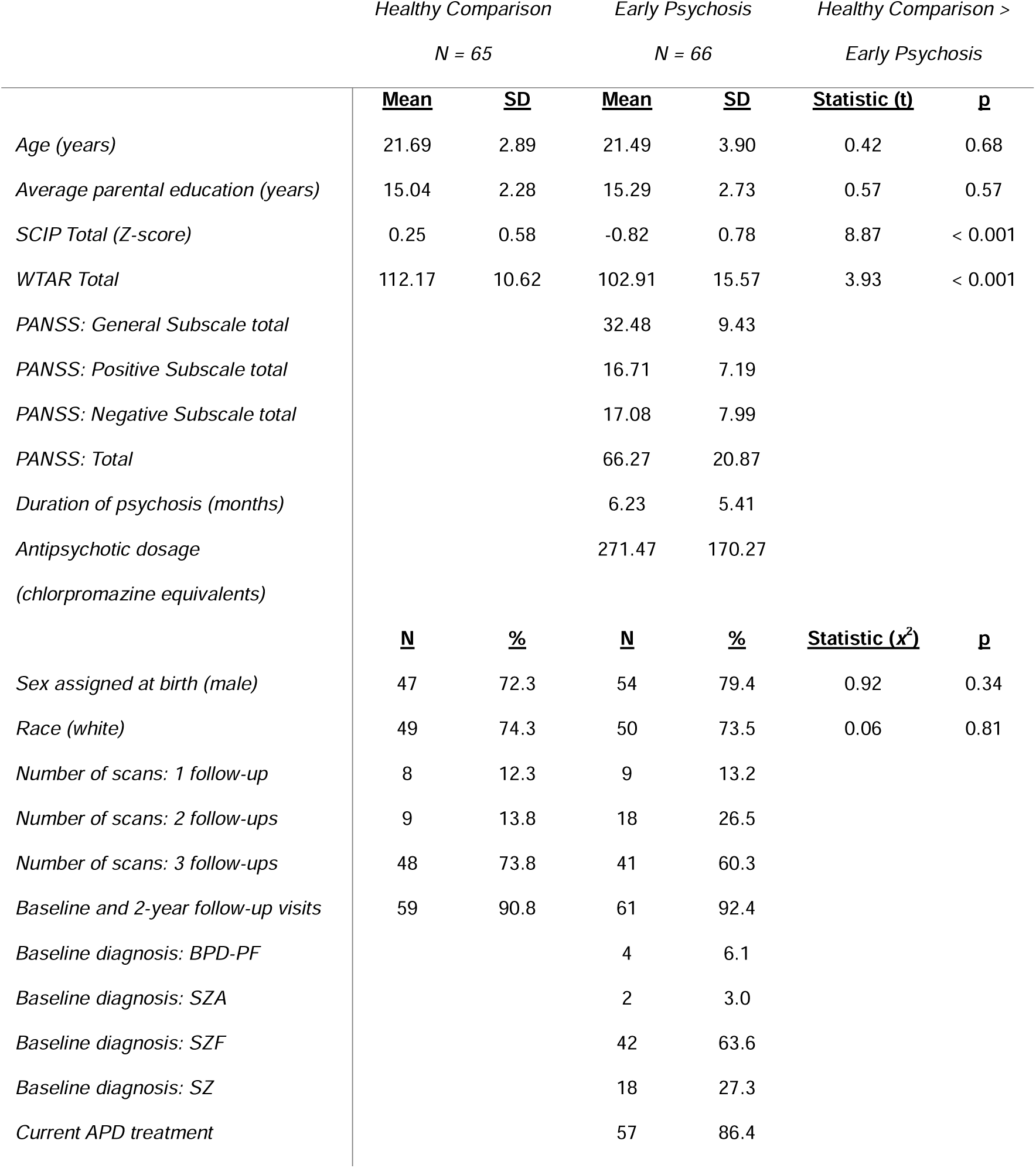
Participant demographics and clinical characteristics at baseline. Abbreviations: SCIP = Screen of Cognition in Psychiatry; WTAR = Wechsler Test of Adult Reading; PANSS = Positive and Negative Syndrome Scale; BPD-PF = bipolar disorder with psychotic features; SZA = schizoaffective disorder; SZF = schizophreniform; SZ = schizophrenia; APD = antipsychotic drug.

From the total sample recruited, 65 HC (90.3%) and 66 EP (89.2%) participants completed scans that passed quality control thresholds. The majority (75% of HC participants and 62.1% of EP participants) of these study participants completed four study visits every 6-8 months over two years, each consisting of a structural MRI scan. Every participant included in analyses completed at least two study visits (Table 1). All participants provided informed consent to be enrolled in the study and were monetarily compensated for each visit they completed. This study was approved by the Vanderbilt University Institutional Review Board. Further details about participant attrition are included in the Supplement (see Figure S1).

### Clinical and cognitive characterization

At baseline and final (i.e., 2-year follow-up) study visits, participants completed an in-person interview including the Structured Clinical Interview for DSM-IV, TR (SCID)^41^. Information from available medical records was reviewed and provided a complement to SCID results; all clinical data was reviewed for diagnostic consensus with S.H., practicing psychiatrist. For EP participants, psychotic symptomatology was assessed using the Positive and Negative Syndrome Scale (PANSS)^42^ at each visit. Current antipsychotic treatment and antipsychotic treatment dosages were assessed at every visit and converted into standardized chlorpromazine equivalents^43,44^.

Diagnostic trajectory subgroupings were operationalized based on an EP participant’s diagnoses at baseline and 2-year follow-up visits. Three trajectories of illness were defined based on whether the participant (1) maintained a diagnosis of schizophreniform disorder over the course of the study (StableSZF; N = 17), (2) entered the study with a diagnosis of schizophreniform disorder at baseline but met criteria for schizophrenia or schizoaffective disorder at 2-year follow-up visit (ProgToSZ; N = 31), or (3) initially met criteria for schizophrenia or schizoaffective disorder upon entering the study (StableSZ; N = 18).

The Wechsler Test of Adult Reading (WTAR) was used to estimate premorbid IQ at baseline^45^. Current cognitive function at each visit was assessed using the Screen for Cognitive Impairment in Psychiatry (SCIP)^46^. At baseline, participants completed the Perceptual Aberrations Scale (PAS), a 35-item True/False validated self-report measure of distortions in perception of the body and other objects^47^. The PAS is composed of statements about psychotic-like bodily distortions and perceptual experiences (e.g., “I can remember when it seemed as though one of my limbs took on an unusual shape.”); changes in body boundaries, parts, or appearance; and unreality (e.g., “I have sometimes felt confused as to whether my body was really my own.”).

More information on clinical and cognitive characteristics across participant groups can be found in Table 1; demographic information for diagnostic trajectory subgroups is located in the Supplement.

### Data acquisition and processing

Structural brain imaging data were collected on one of two 3 Tesla scanners (Philips Healthcare Inc., Best, Netherlands) with 32-channel head coils at the Vanderbilt University Institute of Imaging Sciences in Nashville, TN. At each scanning session, T1-weighted images were collected (voxel size = 1 mm^3^, field of view = 256 mm^2^, number of slices = 170, gap = 0 mm, echo time = 3.7 ms, repetition time = 8.0 ms). All images were visually inspected for artifacts prior to inclusion in analyses; no images were removed. In sum, 458 total images were collected (223 EP, 235 HC)—131 at scan 1, 131 at scan 2, 110 at scan 3, and 86 at scan 4.

T1-weighted images were processed using the CAT12 longitudinal toolbox (Computational Anatomy Toolbox, Structural Brain Mapping group, Jena, Germany)^48^ for voxel-based morphometric analyses of normalized, modulated gray matter volumes. This pipeline is designed specifically for the analysis of longitudinal imaging data and includes spatial registration, tissue classification, intensity bias correction, and scaling. To test our hypotheses that insula subregional volumes change during the early course of psychosis, we used insular subregional masks developed by Farb and colleagues^49^ (see Figure S1) to estimate gray matter volume within the insula. These masks were based on the manual segmentation of insular cortex into agranular, dysgranular, and granular subregions, based on cytoarchitectural characteristics. Briefly, insula subregions of interest (ROIs) were manually drawn by Farb and colleagues to fit each visible gyrus of the template brain in MRIcron, yielding eight anatomically defined regions: five dorsal (four dysgranular and one granular) and three ventral (one agranular, one dysgranular, and one granular). ROIs were collapsed across dorsal and ventral divisions to create three nonoverlapping anatomic masks for insula subregions: agranular, dysgranular, and granular. A whole insula mask was produced by combining these three masks in MRIcron^50^. Subregional volumes were averaged across both hemispheres to reduce parameters in subsequent statistical modeling.

### Statistical analysis

Statistical analyses were carried out using linear mixed effects models in R (R Core Team, 2019) with the lme4, rstatix, emmeans, car, and tidyverse packages^51–55^. A power analysis conducted prior to data analysis determined that with the present sample size, there would be 80% power to detect an effect size of *d*lJ=lJ0.46 in the primary longitudinal analysis with a type 1 error rate of 0.05. Linear mixed effects models testing group differences between HC and EP participants included age at baseline visit, sex assigned at birth, total intracranial volume (TIV), and scanner as fixed effects and included participant as a random effect. Models comparing longitudinal volume differences among diagnostic trajectory subgroups (HC, StableSZF, ProgToSZ, and StableSZ) were constructed using the same fixed and random effects. Cohen’s *d* effect sizes for follow-up contrasts were calculated from the F-statistics using the effectsize package in R^56^. First, to investigate how group influenced insula volumes, we constructed four models with insula Volume (whole, agranular, dysgranular, and granular) as the dependent variables and Group (EP, HC) as a fixed effect. Next, to determine trajectories of insula volume deficits over time in early psychosis, we constructed four separate models with insula Volume (whole, agranular, dysgranular, and granular) as the dependent variable, Group (EP, HC) and Time (months since baseline scan) as fixed effects, and a Group x Time interaction term.

Models were tested with and without interaction terms; main effects reported are from models without interactions. We also tested our hypothesis that subregional insula volumes changed in group-specific ways over the early course of psychosis by constructing one model with Volume as the dependent variable, and Group (EP, HC), Subregion (agranular, dysgranular, and granular), and Time (months since baseline scan) as fixed effects. We then constructed similar models as above, but with Group defined by four diagnostic trajectory subgroups (HC; StableSZF; ProgToSZ; and StableSZ).

Finally, to examine potential brain-behavior relationships between perceptual aberrations and insula volumes, we looked at PAS scores between Group (EP, HC) and tested associations between PAS score and both whole insula volumes and granular insula volumes. We tested these associations across all participants and then evaluated the Group difference. One EP outlier who scored 4+ standard deviations above the mean on the PAS was removed for these analyses. Relationships between insula subregional volumes and clinical and cognitive characteristics in the EP sample were also explored and are provided in the Supplement (see Table S2).

## Results

### Overall differences in insula volumes

Volume was significantly lower in EP participants than HC participants for the whole insula (main effect of Group: F_1,166.97_ = 18.75, p < 0.001, *d* = 0.67; Figure 1) and within all three subregions: agranular (main effect of Group: F_1,128.56_ = 12.02, p < 0.001, *d* = 0.61), dysgranular (main effect of Group: F_1,165.41_ = 16.06, p < 0.001, *d* = 0.62) and granular (main effect of Group: F_1,164.62_ = 14.07, p < 0.001, *d* = 0.58; Figure 1). However, the magnitude of these group differences did not significantly differ across subregions (Group x Subregion interaction: F_3,1691.84_ = 0.49, p = 0.69).

**Figure 1.**
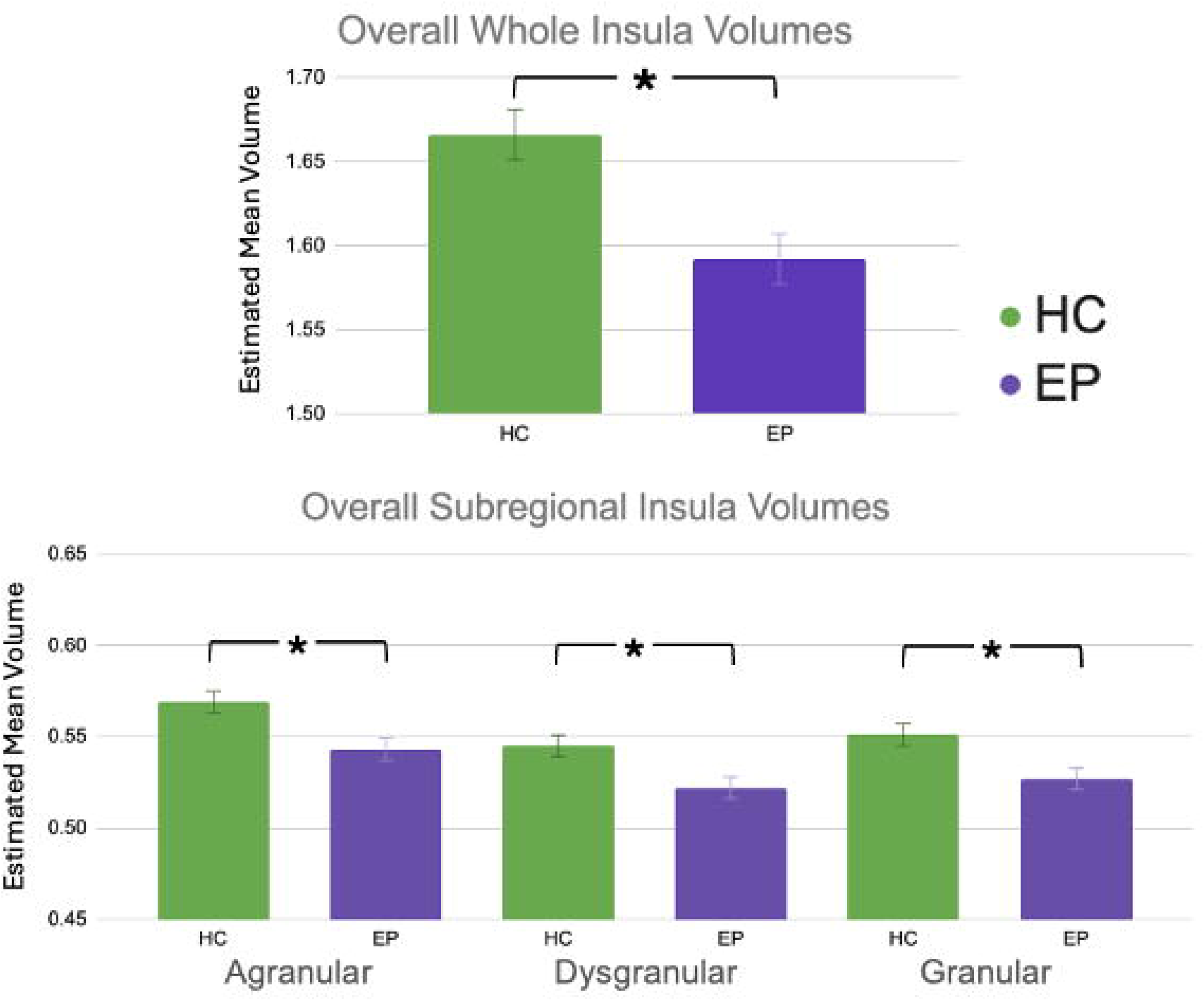
Overall whole, agranular, dysgranular, and granular insula volumes are all lower in EP participants than in HC participants. Error bars represent standard error above and below group means. Abbreviations: HC = healthy comparison; EP = early psychosis. Asterisk (**\***) = p < 0.05.

### Longitudinal differences in insula volumes

In longitudinal models of the whole insula, we found evidence of significantly decreasing insula volumes over two years (main effect of Time: F_1,355.71_ = 11.47, p < 0.001, *d* = 0.36), which differed between groups (Group x Time interaction: F_1,327.87_ = 6.23, p = 0.01, *d* = 0.28; Figure S3, Supplement). This was driven by HC insula volumes decreasing significantly over time (main effect of Time: whole insula: F_1,179.19_ = 23.46, p < 0.001, *d* = 0.72), which was not observed in EP participants (main effect of Time: whole insula: F_1,171.32_ = 0.11, p = 0.74, *d* = 0.05).

Across all participants, we observed volume decline in all insula subregions: agranular (main effect of Time: F_1,349.50_ = 16.57, p < 0.001), dysgranular (main effect of Time: F_1,354.66_ = 3.99, p = 0.046), and granular (main effect of Time: F_1,354.12_ = 13.80, p < 0.001). However, there was no significant Time x Subregion interaction (F_2,1235.58_ = 0.11, p = 0.90). A significant Group x Time interaction was found for the dysgranular and granular subregions (dysgranular: F_1,326.96_ = 11.29, p < 0.001; granular: F_1,326.41_ = 4.02, p = 0.046), again driven by significantly decreasing subregional volumes in HC participants but not in EP participants (see Figure S3). However, the three-way interaction (Group x Subregion x Time) was not significant (F_3,1684.96_ = 0.58, p = 0.63), suggesting that the difference in volumetric trajectories between groups was not significantly different across the three subregions.

### Overall differences in insula volumes: effect of diagnostic trajectory

We next examined how insula volumes differed amongst four diagnostic trajectory subgroups—1) HC (N = 65), 2) StableSZF (N = 17), 3) ProgToSZ (N = 31), and 4) StableSZ (N = 18). Whole insula and all subregions significantly differed across these four groups (whole insula main effect of Trajectory: F_3,124.22_ = 5.76, p = 0.001; agranular main effect of Trajectory: F_3,124.82_ = 4.75, p = 0.004; dysgranular main effect of Trajectory: F_3,123.39_ = 4.55, p = 0.005; granular main effect of Trajectory: F_3,122.61_ = 4.18, p = 0.007). We did not find subregional specificity in insula volume deficits that varied with diagnostic trajectory (Trajectory x Subregion interaction: F_6,1231.44_ = 1.12, p = 0.35).

Post-hoc, Bonferroni-corrected pairwise comparisons were conducted to examine the pattern of volume differences across trajectory groups (Table 2). HC participants had significantly greater insula volumes across all measures compared only to the stable schizophrenia subgroup (main effect of Trajectory for HC vs. StableSZ: whole insula: F_1,82.24_ = 12.46; p_corrected_ < 0.001, *d* = 0.78; agranular insula: F_1,81.54_ = 10.32; p_corrected_ = 0.01, *d* = 0.71; dysgranular insula: F_1,81.39_ = 9.82; p_corrected_ = 0.01, *d* = 0.69; granular insula: F_1,80.97_ = 9.18; p_corrected_ = 0.02, *d* = 0.67; Figure 2). None of the diagnostic trajectory groups differed from one another. Of note, duration of illness did not significantly correlate with volume for any of the subregions (see Table S2).

**Table 2.** Comparisons of insula volumes (whole, agranular, dysgranular, and granular) for each diagnostic trajectory subgroup pairing. Significance was determined at p < 0.008 for six comparisons; p values shown below are Bonferroni-uncorrected. Significant comparisons that survived Bonferroni correction for multiple comparisons are **bolded**. Abbreviations: SZF = schizophreniform; SZ = schizophrenia.

| *Trajectory Pairing* | *Whole Insula*<br>*Volume* | *Agranular*<br>*Subregional Volume* | *Dysgranular*<br>*Subregional Volume* | *Granular*<br>*Subregional Volume* |
| --- | --- | --- | --- | --- |
| *HC-StableSZF* | $F_{1,76.53} = 7.44$;<br>$p = 0.008$ | $F_{1,77.49} = 6.25$;<br>$p = 0.014$ | $F_{1,75.78} = 4.34$;<br>$p = 0.041$ | $F_{1,75.56} = 7.05$;<br>$p = 0.001$ |
| *HC-ProgToSZ* | $F_{1,92.03} = 3.80$;<br>$p = 0.05$ | $F_{1,91.71} = 3.93$;<br>$p = 0.05$ | $F_{1,91.73} = 1.97$;<br>$p = 0.16$ | $F_{1,91.24} = 3.07$;<br>$p = 0.08$ |
| *HC-StableSZ* | **$F_{1,82.24} = 12.46$;**<br>**$p < 0.001$** | **$F_{1,81.54} = 10.32$;**<br>**$p = 0.002$** | **$F_{1,81.39} = 9.82$;**<br>**$p = 0.002$** | **$F_{1,80.97} = 9.18$;**<br>**$p = 0.003$** |
| *StableSZF-ProgToSZ* | $F_{1,39.35} = 0.80$;<br>$p = 0.38$ | $F_{1,41.07} = 0.10$;<br>$p = 0.75$ | $F_{1,37.63} = 1.32$;<br>$p = 0.25$ | $F_{1,40.65} = 0.75$;<br>$p = 0.39$ |
| *StableSZF-StableSZ* | $F_{1,28.54} = 0.33$;<br>$p = 0.57$ | $F_{1,28.81} = 0.10$;<br>$p = 0.76$ | $F_{1,28.25} = 0.31$;<br>$p = 0.58$ | $F_{1,29.42} = 0.45$;<br>$p = 0.51$ |
| *ProgToSZ-StableSZ* | $F_{1,44.73} = 2.27$;<br>$p = 0.14$ | $F_{1,44.11} = 1.04$;<br>$p = 0.31$ | $F_{1,44.89} = 3.66$;<br>$p = 0.062$ | $F_{1,44.33} = 1.19$;<br>$p = 0.28$ |

**Figure 2.**
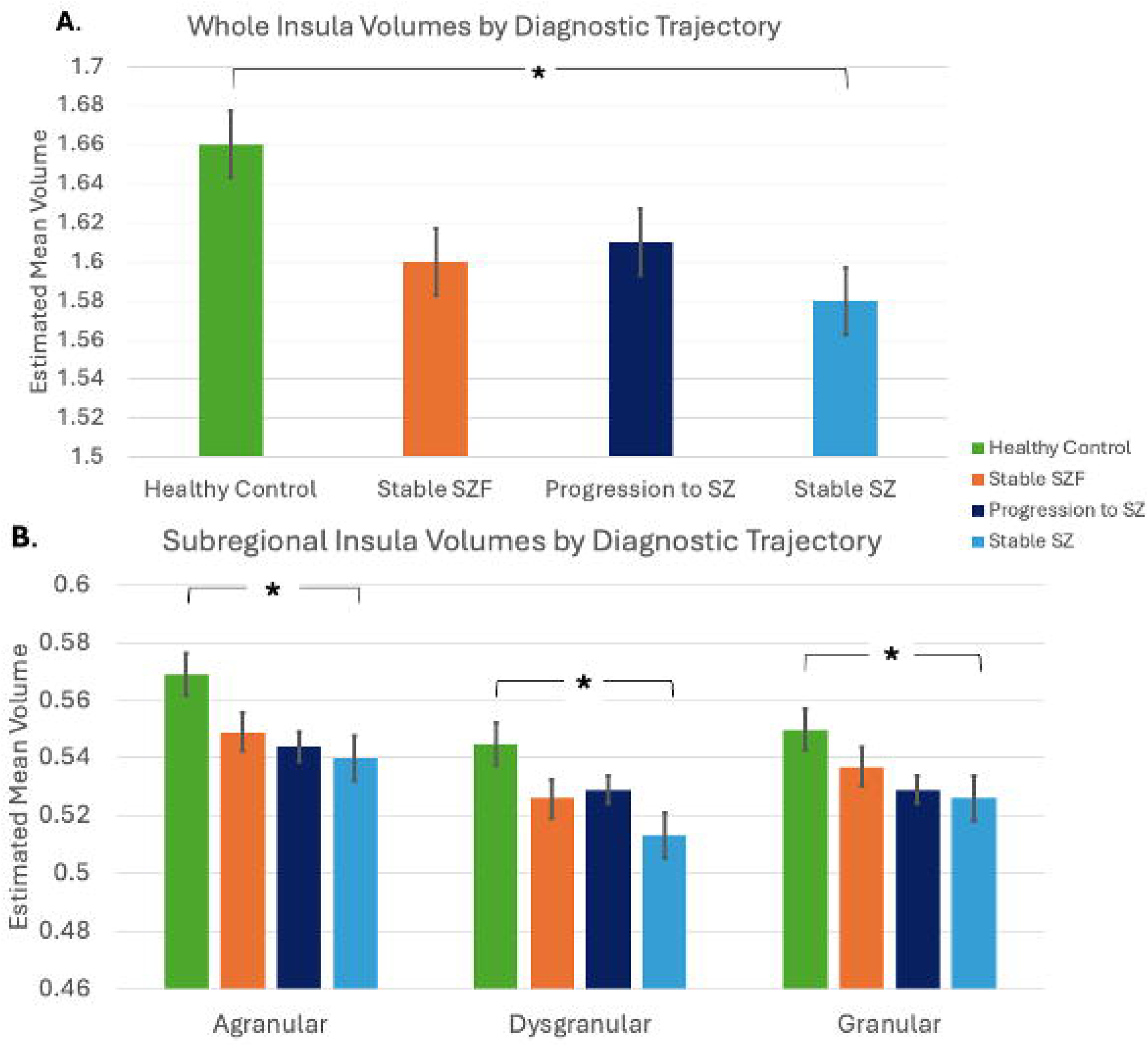
Whole insula volumes differed significantly among diagnostic trajectory subgroups (**A**), as did agranular, dysgranular, and granular subregional insula volumes (**B**). Significant pairwise differences (Bonferroni-corrected p < 0.008 for six comparisons) are indicated for each volume measure by **\***; error bars represent standard error above and below subgroup estimated marginal means. Abbreviations: SZF = schizophreniform; SZ = schizophrenia. Asterisk (**\***) = p < 0.05.

### Longitudinal differences in insula volumes: effect of diagnostic trajectory

We did not find evidence of differences in whole insula volume change over time, based on diagnostic trajectory (Trajectory x Time interaction: F_3,323.91_ = 2.38, p = 0.07; Figure 3), nor did we find any subregional specificity in insula volume deficits that varied with diagnostic trajectory over time (Trajectory x Subregion x Time interaction: F_6,1220.54_ = 0.42, p = 0.86). Follow-up analyses revealed no evidence of significant changes in any insula volume (whole, agranular, dysgranular, or granular) over time in any of the three clinical trajectory subgroups (Table 3).

**Figure 3.**
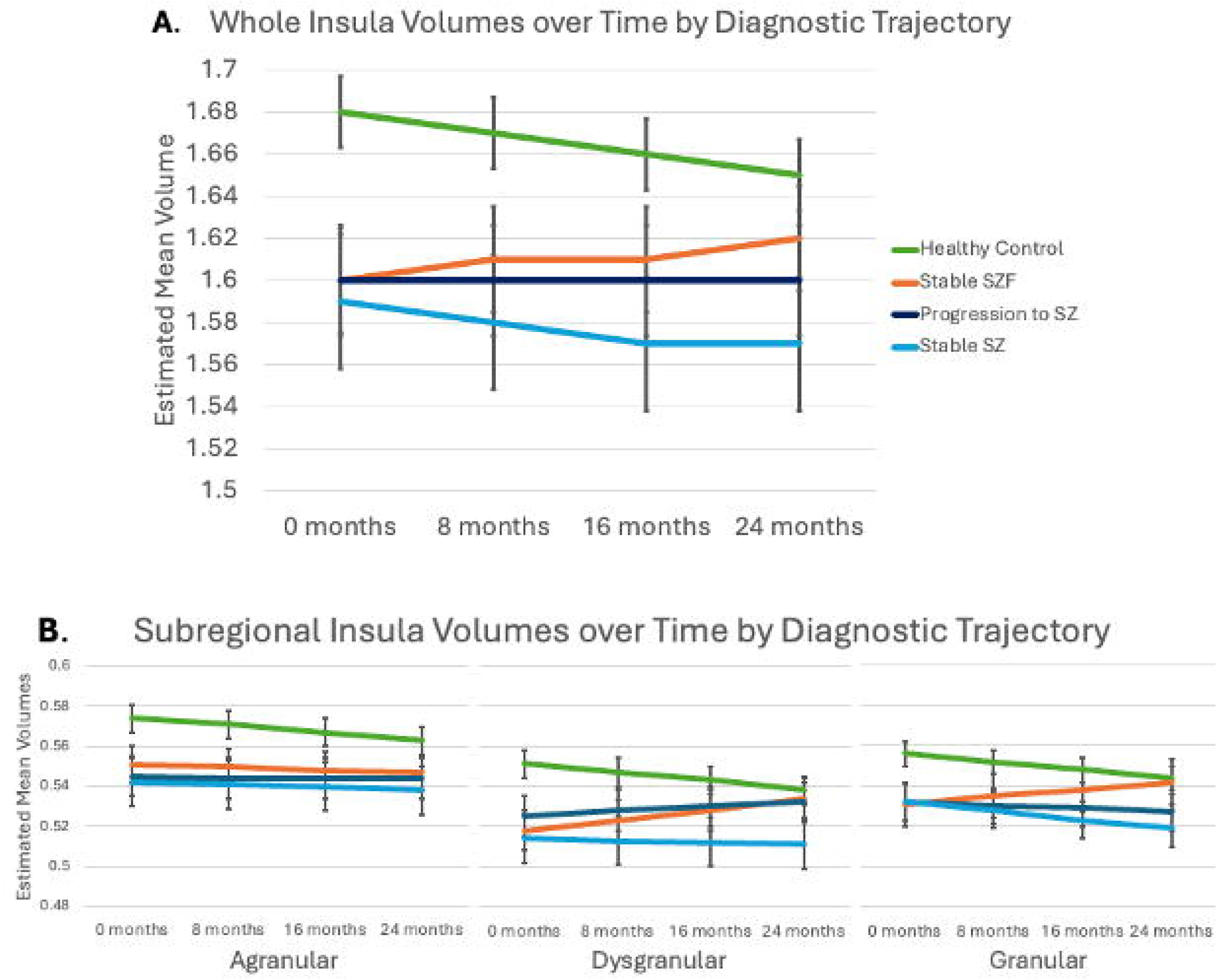
Whole (**A**) insula volumes and some (dysgranular and granular) but not all subregional (**B**) insula volumes change differentially over time among diagnostic trajectory subgroups. Volumes shown are estimated marginal means at timepoints 0 months, 8 months, 16 months, and 24 months; error bars represent standard error above and below the estimated marginal mean. Abbreviations: SZF = schizophreniform disorder; SZ = schizophrenia.

**Table 3.** Main effect of time for each diagnostic trajectory subgroup for each insula volume measure. Bonferroni-corrected p < 0.013 are considered significant (four comparisons per volume measure). Significant comparisons that survived Bonferroni correction for multiple comparisons are **bolded**. Abbreviations: SZF = schizophreniform; SZ = schizophrenia.

| *Trajectory* | *Whole Insula* | *Agranular* | *Dysgranular* | *Granular* |
| --- | --- | --- | --- | --- |
| *Subgroup* | *Volume* | *Subregional Volume* | *Subregional Volume* | *Subregional Volume* |
| **HC** | **F<sub>1,179.19</sub> = 23.46;**<br>**p < 0.001** | **F<sub>1,176.20</sub> = 22.55;**<br>**p < 0.001** | **F<sub>1,177.68</sub> = 17.45;**<br>**p < 0.001** | **F<sub>1,180.37</sub> = 20.62;**<br>**p < 0.001** |
| **StableSZF** | F<sub>1,39.41</sub> = 0.09;<br>p = 0.76 | F<sub>1,40.25</sub> = 3.18;<br>p = 0.082 | F<sub>1,38.49</sub> = 0.15;<br>p = 0.70 | F<sub>1,41.03</sub> = 0.03;<br>p = 0.86 |
| **ProgToSZ** | F<sub>1,74.92</sub> = 0.83;<br>p = 0.37 | F<sub>1,72.99</sub> = 0.25;<br>p = 0.62 | F<sub>1,76.49</sub> = 2.72;<br>p = 0.10 | F<sub>1,73.51</sub> = 0.04;<br>p = 0.84 |
| **StableSZ** | F<sub>1,46.58</sub> = 0.66;<br>p = 0.42 | F<sub>1,45.52</sub> = 0.49;<br>p = 0.49 | F<sub>1,46.29</sub> = 0.08;<br>p = 0.78 | F<sub>1,45.92</sub> = 1.95;<br>p = 0.17 |

### Baseline associations of insula volumes with perceptual aberrations

PAS scores were significantly elevated in EP compared to HC participants at baseline (*H* = 23.32, p < 0.001; Figure 4A). Across all participants, we did not find evidence for a significant association between PAS total and baseline whole insula volume (r_S_ = -0.06, p = 0.50; Figure 4B) or baseline granular volume (r_S_ = -0.05, p = 0.58; Figure 4C). Finally, based on our a priori interest in the granular insula, we did not find evidence of baseline PAS total scores significantly predicting granular subregional volume changes across EP participants in a linear mixed model (main effect of PAS: F_1,58.69_ = 0.026, p = 0.87).

**Figure 4.**
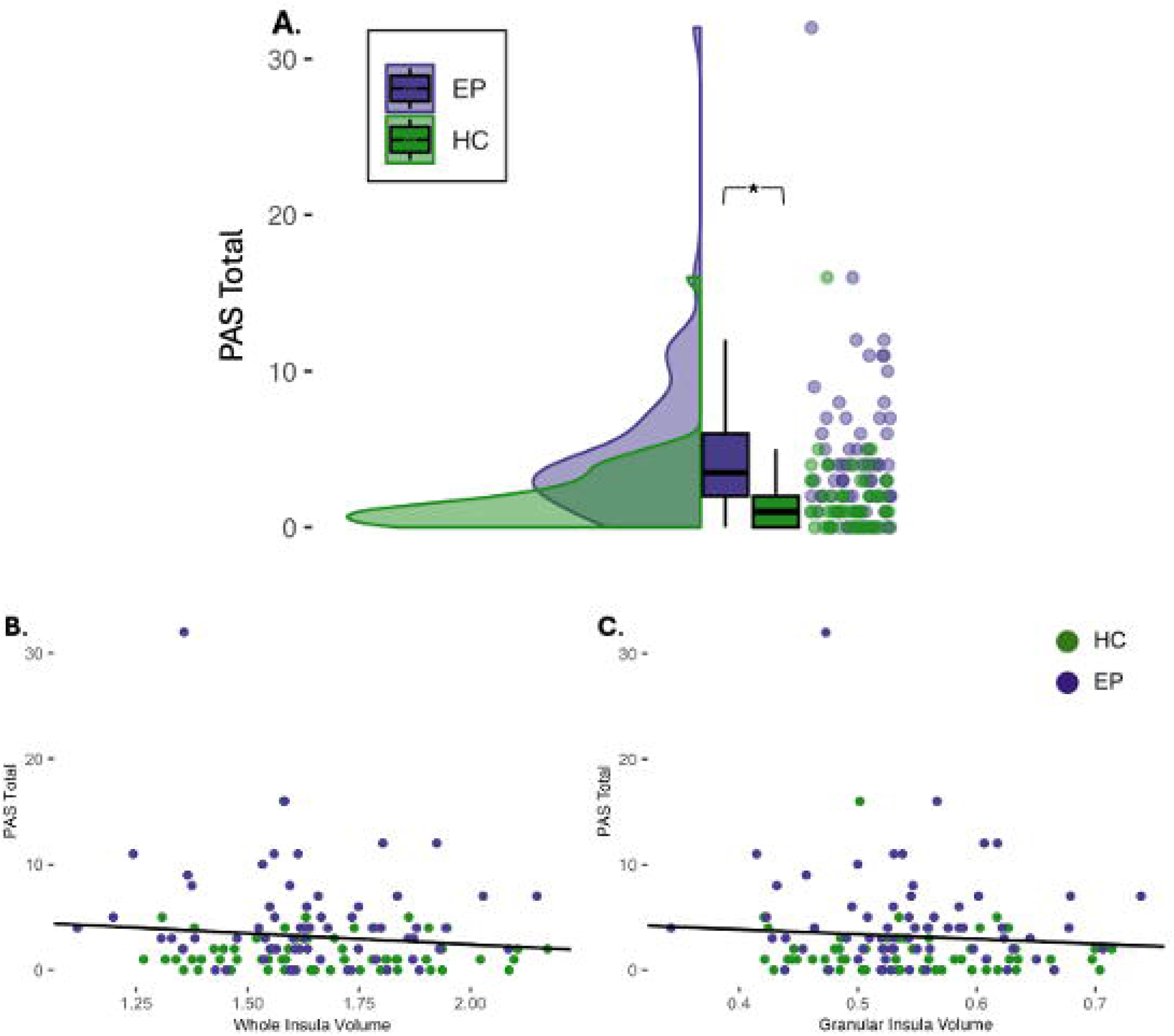
Self-reported perceptual aberrations (as measured by PAS total) are higher in EP participants at baseline (**A**). Neither whole insula volumes (**B**) nor granular insula subregional volumes (**C**) were significantly associated with self-reported perceptual aberrations at baseline among all participants. Abbreviations: EP = early psychosis; HC = healthy comparison; PAS = Perceptual Aberrations Scale. Asterisk (**\***) = p < 0.05.

## Discussion

With a longitudinal case-control study design we show that insula volumes are smaller by first episode of psychosis and do not change over two years. This baseline deficit is present in all granular subregions without apparent specificity, although individuals who already meet criteria for schizophrenia at the time of study entry show evidence of the largest deficits compared to nonclinical samples. Finally, despite higher self-reported perceptual aberrations in EP participants, no relationships were observed with insula volumes. Overall, our data demonstrate that individuals with schizophrenia-spectrum disorders have smaller insula volumes across all granular subregions at first episode of psychosis and that trajectory of illness may be important for understanding this process. Our results suggest that structural abnormalities of the insula occur prior to illness onset as neurodevelopmental insults.

The development of the insula as a cortical brain region consists of several overlapping but temporally distinct processes, beginning with gyrification in prenatal development^57,58^, followed by sulcal deepening soon after birth^59,60^, cortical thickening in early childhood^61,62^, and subsequent cortical thinning in adolescence into early adulthood^63^. Prior work has shown perturbations to the insula in all of these structural metrics in individuals with psychosis^64–68^.

Excessive synaptic pruning during adolescence has long been proposed as one mechanism by which individuals with schizophrenia develop disproportionally high gray matter loss^69^, and neuroimaging studies of adolescents with child-onset psychosis have shown evidence of earlier and more severe cortical gray matter loss compared to non-clinical samples^70^. Importantly, there seems to be some diagnostic specificity in these perturbations, as similar structural alterations to the insula have not been observed in individuals with other affective psychotic disorders such as psychotic bipolar disorder^28^. These morphological abnormalities in the insula, seemingly unique to schizophrenia, may underlie the earliest phenomenological signs of illness.

Furthermore, it is important to note that our individuals with psychosis did not demonstrate gradually declining volumes over time as HC participants did. This may be due to prior gray matter loss in the prodrome. Indeed, across individuals at clinical high-risk for psychosis, cortical gray matter volume declines steepest for those who will convert to psychosis^71^, including within the insula^10^. In contrast, in studies of individuals with established schizophrenia-spectrum disorders, insula volumes did not decline progressively over time^72^. Taken together, this literature on insula structure in psychosis suggests that volume changes predate diagnosis onset, occur at least in part during the prodrome, and subsequently lessen in more chronic stages. The results of this study situate well into this existing literature by confirming that insula structural changes in psychosis occur prior to the onset of illness and may be pronounced for individuals with more severe illness trajectories, reflected by the smallest insula volumes among individuals who already met criteria for schizophrenia at the time of the study.

In addition to the neurobiological mechanism underlying these structural brain changes in psychosis, it is crucial to better understand how the resulting aberrant volumes may affect clinical phenotypes, and how the insula may impact psychotic phenomenology more broadly. With its varied roles in somatosensation, salience detection, and social cognition, the insula is a key brain region for understanding the phenomenology of reality distortions, perceptual aberrations, and social cognitive impairments that characterize psychosis^39,73,74^. Prior literature has proposed that insula subregions are important for specific realms of function in normative populations and dysfunction in schizophrenia^16,32^. Thus, the extent of volume loss in specific insula subregions may also provide a neural basis for worsening psychotic symptomatology in specific domains, which has been shown across various realms of function and subregions of the insula^75^. Our evidence of comparable volume loss across all three subregions of the insula in early psychosis suggests that all associated symptom realms may be affected.

With this context, we explored how insula gray matter volume might be associated with self-reported perceptual aberrations to investigate other brain-behavior relationships relevant to the insula. The insula is a key node within a proposed “interoceptive network” in schizophrenia^76^, so we investigated how volume changes might affect interoceptive deficits to extend our imaging findings into clinically meaningful brain-behavior relationships. A previous study by Brosey and Woodward of perceptual aberrations and insula volumes found a significant negative correlation between PAS scores and bilateral whole and granular insula volumes in participants with chronic psychosis^39^, which was not observed in our cohort. In fact, while our EP participants did score higher on the PAS than HC participants and recapitulate prior behavioral findings^77^, we did not find evidence of any significant relationships between self-reported perceptual aberrations at baseline and insula volumes. However, it may be that this association is not apparent until symptoms progress further; our cohort was captured soon after illness onset, while Brosey and Woodward’s cohort was older and more chronic in their illness course.

Additionally, the phenomenology of psychosis is highly complex and multifaceted, requiring many brain regions and circuits to coordinate efforts^78^. Indeed, perceptual aberrations represent one such behavioral readout of psychotic symptomatology, and multiple brain regions, including but not limited to the insula, are involved in their production^39^. Probing the functional activation and connectivity of the insula, instead of investigating structural changes as here, may in fact be better for understanding links between this region and symptoms of schizophrenia-spectrum disorders.

In sum, the current study is the first longitudinal investigation of both insula subregional volume and diagnostic trajectories in the early period of psychosis, and provides a valuable view into brain changes during this critical window. Our study suggests that insula volume changes in early psychosis differ from typical neurodevelopmental trajectories, and that illness-related volume changes likely precede clinical presentation and diagnosis. We also provide evidence here that among those with an emerging psychotic disorder, insula volumes are most different among patients with chronic schizophrenia compared to nonclinical comparisons. This study provides insight into the underlying neurobiological changes in the insula that may precede or occur alongside symptom progression in psychotic illness. Altered insula volumes, especially at the subregional level, may be an important developmental factor among those at high risk for developing schizophrenia.

## Data Availability

All data produced in the present study are available upon reasonable request to the authors.

## Acknowledgments

The work outlined in this manuscript was supported by the Charlotte and Donald Test Fund, NIMH grants R01-MH70560 (Heckers) R01-MH123563 (Vandekar), the Jack Martin, MD Research in Psychopharmacology endowment (Sheffield) and K23-MH126313. This work was conducted in part using the resources of the Center for Computational Imaging at the Vanderbilt University Institute of Imaging Sciences and the Advanced Computing Center for Research and Education (ACCRE) at Vanderbilt University in Nashville, TN. The authors would like to thank the participants of this study for their involvement in and commitment to research.

