## Supplement for "A 2-year longitudinal investigation of insula subregional volumes in early psychosis"

**Supplementary Methods**

*Inclusion and exclusion criteria*

Inclusion criteria for all participants were (1) age 13-40 years and (2) estimated premorbid IQ greater than or equal to 75 via the Wechsler Test of Adult Reading (WTAR)^44^; additional inclusion criteria for early psychosis (EP) participants were (3) less than two years of psychosis and (4) meeting criterion A for schizophrenia from DSM-IV^75^, with at least one month’s duration of two of the following: hallucinations, delusions, disorganized speech, disorganized or catatonic behavior, and negative symptoms. Additionally, at two-year follow-up, only EP participants who met criteria for schizophreniform disorder, schizoaffective disorder, or schizophrenia were included for longitudinal analyses, which allowed us to create diagnostic trajectory subgroups based on follow-up diagnosis for exploratory analyses. Exclusionary criteria for all participants included head injury with loss of consciousness, significant medical illness, or pregnancy. Additional exclusion criteria for EP participants also included meeting criteria for substance use disorder within the last month and being diagnosed with psychosis secondary to a medical condition, while additional exclusion criteria for healthy comparison (HC) participants also included any Axis I psychiatric diagnosis and family history of first-degree relative with a diagnosed psychotic disorder.

**Supplementary Results**

*Clinical and cognitive characteristics in the overall sample*

EP and HC participants were matched on all basic demographic variables (age at baseline visit, sex assigned at birth, average parental education, and self-reported race; Table 1, main text). At baseline, EP participants had lower premorbid IQ as estimated by the WTAR (t(132) = 3.93, p < 0.001), lower current cognitive functioning as estimated by the SCIP (t(132) = 8.87, p < 0.001), and higher self-reported perceptual aberrations as measured by the PAS (t(132) = 4.25, p < 0.001).

*Clinical and cognitive characteristics by diagnostic trajectory*

Demographic, clinical, and cognitive characteristics of our diagnostic trajectory subgroups are detailed in Table S1. At baseline, our three clinical subgroups (StableSZF, ProgToSZ, and StableSZ) were matched on age at baseline visit, sex assigned at birth, self-reported race, and average parental education, as well as premorbid and baseline cognitive abilities as measured by the WTAR and SCIP, respectively. Our clinical subgroups differed in symptomatology captured by the PANSS positive subscale (F_2,63_ = 4.12, p = 0.021) but not negative (F_2,63_ = 0.06, p = 0.94) or general (F_2,63_ = 1.01, p = 0.37) subscales, and the subgroups did not significantly differ in their total symptom severity (F_2,63_ = 1.44, p = 0.25). Our diagnostic trajectory subgroups at baseline also did not differ in the number of individuals taking antipsychotic drugs (F_2.63_ = 0.17, p = 0.84), nor in the CPZ equivalents of antipsychotic dosage (F_2,63_ = 1.56, p = 0.22).

**Supplementary Tables**

**Table S1**. Baseline clinical and cognitive characterization of early psychosis participants by diagnostic trajectory subgrouping. Abbreviations: WTAR = Wechsler Test of Adult Reading; SCIP = Screen for Cognitive Impairment in Psychosis; PANSS = Positive and Negative Syndrome Scale; CPZ = chlorpromazine; BPD-PF = bipolar affective disorder with psychotic features; SZA = schizoaffective disorder; SZ = schizophrenia; SZF = schizophreniform disorder; APD = antipsychotic drug.

|  | StableSZF  N = 17 | | ProgToSZ  N = 31 | | StableSZ  N = 18 | |  |  |
| --- | --- | --- | --- | --- | --- | --- | --- | --- |
|  | **Mean** | **SD** | **Mean** | **SD** | **Mean** | **SD** | **Statistic** | **p** |
| Age (years) | 21.12 | 3.48 | 20.97 | 4.33 | 22.56 | 3.50 | F_2,63_ = 1.01 | 0.37 |
| Parental education (years) | 15.85 | 3.20 | 14.92 | 2.35 | 15.42 | 3.01 | F_2,63_ = 0.65 | 0.53 |
| WTAR total | 110.18 | 11.29 | 99.10 | 17.37 | 101.00 | 13.64 | F_2,63_ = 3.10 | 0.052 |
| SCIP Total (Z-score) | -0.59 | 0.62 | -0.81 | 0.92 | -1.01 | 0.62 | F_2,63_ = 1.31 | 0.28 |
| PANSS: Positive | 15.0 | 6.59 | 15.35 | 6.72 | 20.67 | 7.38 | **F_2,63_ = 4.12** | **0.02** |
| PANSS: Negative | 16.53 | 6.56 | 17.16 | 9.05 | 17.44 | 7.66 | F_2,63_ = 0.06 | 0.94 |
| PANSS: General | 31.24 | 8.02 | 31.61 | 10.62 | 35.17 | 8.38 | F_2,63_ = 1.01 | 0.37 |
| PANSS: Total | 62.76 | 17.33 | 64.13 | 23.38 | 73.28 | 18.60 | F_2,63_ = 1.44 | 0.25 |
| Duration of psychosis (months) | 4.94 | 6.64 | 4.58 | 4.16 | 10.61 | 4.02 | **F_2,63_ = 9.62** | **< 0.001** |
| CPZ equivalents | 209.82 | 151.64 | 287.86 | 193.62 | 301.44 | 133.46 | F_2,63_ = 1.56 | 0.22 |
|  | **N** | **%** | **N** | **%** | **N** | **%** | **Statistic** | **p** |
| Sex (male) | 14 | 82.4 | 23 | 74.2 | 15 | 83.3 | F_2,63_ = 0.36 | 0.70 |
| Race (white) | 15 | 88.2 | 24 | 77.4 | 11 | 61.1 | F_2,63_ = 2.09 | 0.13 |
| Number of scans: 1 follow-up* | 3 | 17.6 | 8 | 25.8 | 2 | 11.1 |  |  |
| Number of scans: 2 follow-ups | 3 | 17.6 | 7 | 22.6 | 5 | 27.8 |  |  |
| Number of scans: 3 follow-ups | 11 | 64.7 | 16 | 51.6 | 11 | 61.1 |  |  |
| Baseline diagnosis: BPD-PF | 0 | 0 | 4 | 12.9 | 0 | 0 |  |  |
| Baseline diagnosis: SZA | 0 | 0 | 0 | 0 | 2 | 11.1 |  |  |
| Baseline diagnosis: SZF | 17 | 100 | 27 | 87.1 | 0 | 0 |  |  |
| Baseline diagnosis: SZ | 0 | 0 | 0 | 0 | 16 | 88.9 |  |  |
| Current APD treatment | 14 | 82.4 | 27 | 87.1 | 16 | 88.9 | F_2,63_ = 0.17 | 0.84 |

**Table S2.** Associations of clinical characteristics with baseline mean insula subregional volumes in early psychosis (EP) participants. Among EP participants, there were no significant associations between insula volumes at baseline and any realms of psychotic symptomatology, duration of psychotic illness, or antipsychotic dosage. Abbreviations: PANSS = Positive and Negative Syndrome Scale; CPZ = chlorpromazine.

|  | Agranular Insula Subregion | | Dysgranular Insula Subregion | | Granular Insula Subregion | |
| --- | --- | --- | --- | --- | --- | --- |
| Clinical Characteristic | Statistic (r) | p | Statistic (r) | p | Statistic (r) | p |
| PANSS Total Score | -0.02 | 0.88 | -0.10 | 0.43 | -0.14 | 0.26 |
| Positive Subscale | 0.06 | 0.63 | -0.05 | 0.68 | -0.10 | 0.44 |
| Negative Subscale | 0.04 | 0.72 | 0.02 | 0.89 | -0.03 | 0.79 |
| General Subscale | -0.12 | 0.34 | -0.19 | 0.12 | -0.21 | 0.09 |
| Duration of psychosis (months) | -0.08 | 0.53 | -0.14 | 0.25 | -0.10 | 0.43 |
| Antipsychotic dosage (CPZ equivalents) | 0.05 | 0.70 | 0.04 | 0.73 | 0.05 | 0.69 |

**Supplementary Figures**

**Figure S1**. Study participant attrition. Abbreviations: EP = early psychosis.

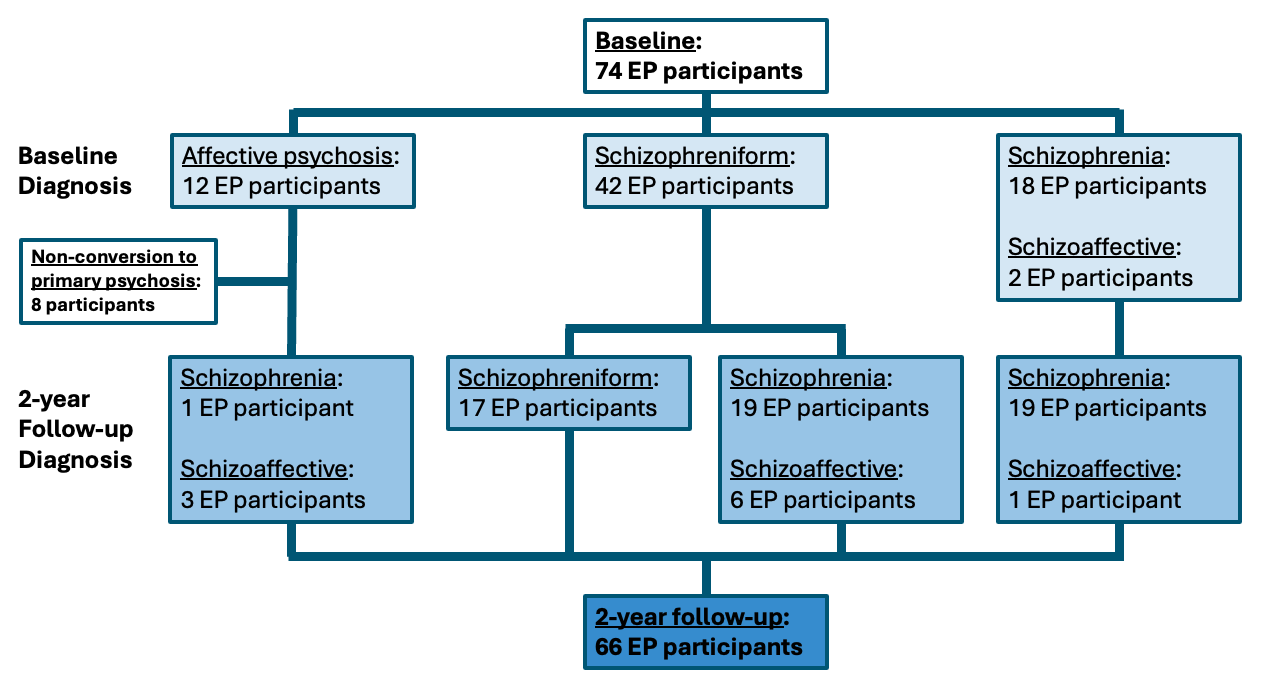

**Figure S2.** Insula subregions were manually drawn to create three non-overlapping anatomic masks for insula subregions: agranular (green), dysgranular (blue), and granular (red). Slice shown at coordinates x = 39, y = 9, z = 1.

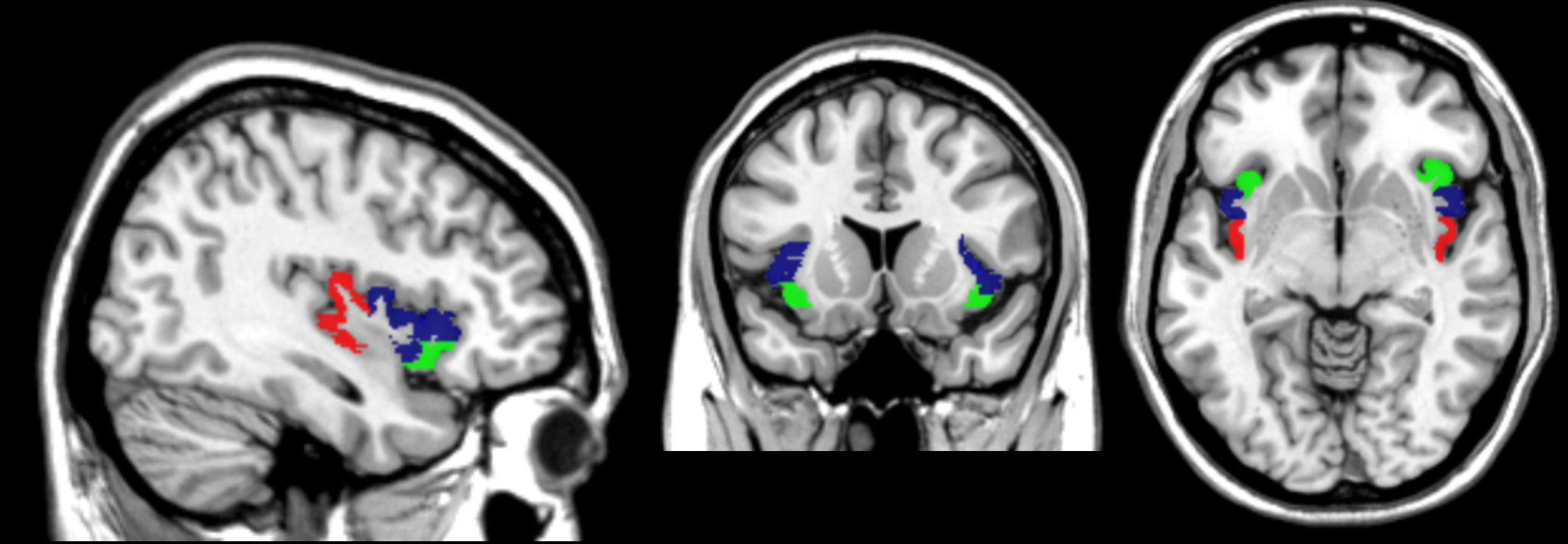

**Figure S3.** Insula volume is reduced in the whole insula as well as the dysgranular and granular subregions in early psychosis (EP) participants compared to healthy comparisons (HC). Significantly different patterns of change in volume over time are driven by stable volumes in EP participants and declining volumes in HC participants. Error bars indicate standard error above and below the estimated marginal mean at times 0, 8, 16, and 24 months. Abbreviations: HC = healthy comparison; EP = early psychosis.

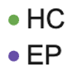

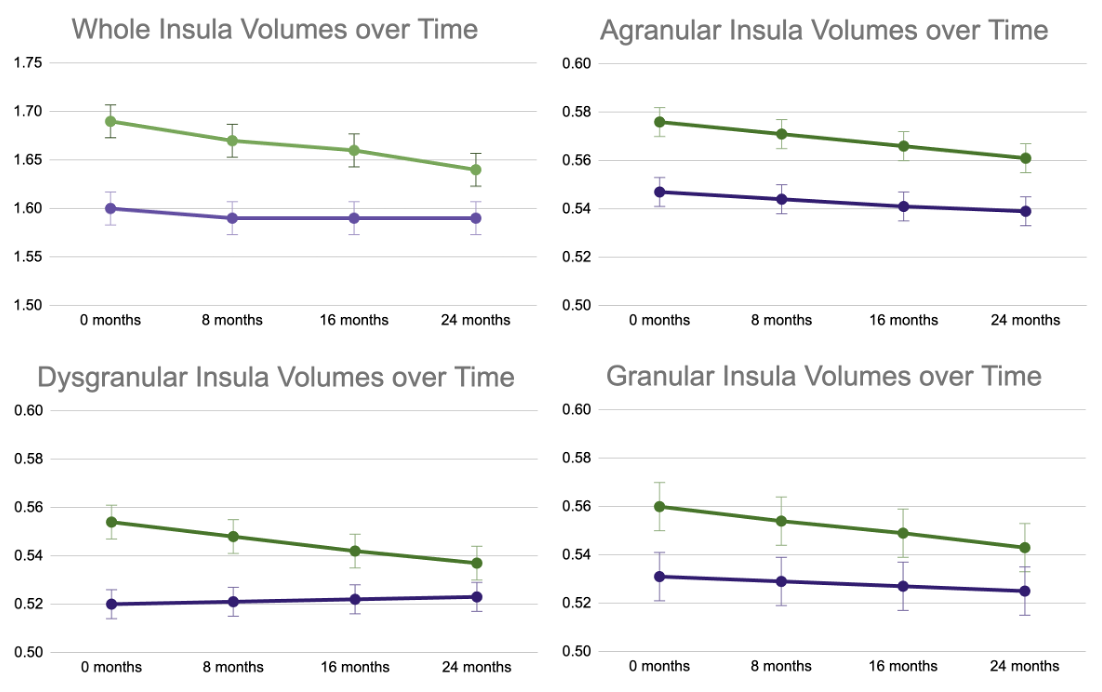

Estimated Mean Volumes

Estimated Mean Volumes
